# The Impact of Hurricane Helene on the Incidence of Acute Gastrointestinal Illness in North Carolina, US: A Controlled Interrupted Time Series Analysis

**DOI:** 10.64898/2026.11.18.26368308

**Authors:** Jahred M. Liddie, Katherine R. O’Brien, Amina Pasha, Abigail Iseson, Tatiyana V. Apanasovich, Pramita Bagchi, Agnes Pastwa, Xindi C. Hu

**Author notes:** Correspondence to: Jahred Liddie, Department of Environmental and Occupational Health, George Washington Milken Institute School of Public Health, 950 New Hampshire Ave NW #2, Washington, D.C. 20037.

## Abstract

In September 2024, Hurricane Helene caused significant flooding in western North Carolina (NC), leading to concerns surrounding water quality, including the risk of acute gastrointestinal illness (AGI). The potential for flooding-related drinking water contamination is exacerbated for private well users, who comprised a large proportion of the population most affected by the hurricane. Here we investigated the impact of Hurricane Helene on AGI and determined if this impact was different in areas with greater proportions of residents using private wells. We used electronic health records from Truveta to identify inpatient and outpatient visits to hospitals for AGI in NC from 2016-2024 at the zip3-by-week level and classified exposure to flooding using satellite-derived flood depth estimates. We conducted a controlled interrupted time series analysis to estimate the effect of the hurricane on the incidence of AGI among flooded zip3 units. From 2017 to 2024, there were approximately 548k AGI visits in NC across 20 zip3 units. Our findings show that the hurricane was associated with positive, insignificant changes in the incidence of AGI three weeks following the hurricane (incidence rate ratio: 1.11 [0.59, 2.07]), controlled for pre-hurricane trends and post-hurricane changes in lesser flooded regions. Results were consistent across several time windows and analyses examining sensitivity to geographic data masking, model specification, and study design. Our findings warrant re-evaluation at finer spatial and temporal scales and examination of other public health impacts of hurricanes, which are expected to increase in severity and frequency in the eastern United States due to climate change.

**Graphical abstract:** 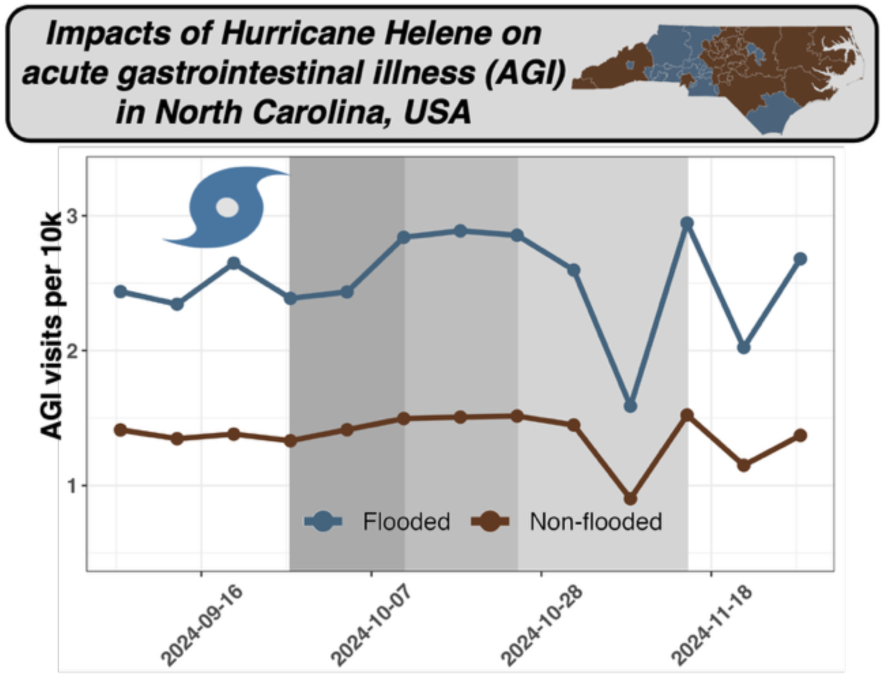

## 1. Introduction

Hurricanes are expected to increase in both frequency and intensity due to climate change (1). These events have widespread impacts on health, including but not limited to physical injuries, waterborne diseases, and mental health issues (2). The disruption of health care services and public health systems, which need to shift from normal operations to emergency response, further compounds the impacts of these events (1,3). The impact of hurricanes can be significant for drinking water systems, particularly individuals using private wells (4). Over 43 million United States (US) residents (13% of the population), many in rural or underserved areas, rely on private wells for drinking water (5). Because private wells are unregulated by the federal government, water contamination can often remain undetected unless well users conduct water testing independently.

From 1980-2024, North Carolina (NC) has experienced 33 tropical cyclones and flooding events with financial losses exceeding 1 billion USD. In September 2024, Hurricane Helene caused widespread destruction in rural, western NC and is considered the most devastating disaster in the region’s history (6). Rainfall amounts ranged from 20-30 inches, and the state experienced severe flooding and landslides, resulting in 108 deaths (6,7). Counties in declared disaster zones following the hurricane faced barriers to healthcare access and generally had greater proportions of individuals with disabilities, older adults, and mobile homeowners (8,9). The 25 western counties of North Carolina that were most affected by Hurricane Helene are also home to many rural communities relying on private wells, which can have elevated risk of contamination and are not subject to routine monitoring (10). Across all of NC, roughly 25% of residents rely on private wells and only 6% of the predicted 1.6 million wells have been tested since February 2009 (11,12), leading to significant knowledge gaps in water quality for many individuals even in the absence of extreme weather conditions.

Acute gastrointestinal illness (AGI) broadly describes enteric illness caused by *Escherichia coli, Salmonella, Campylobacter, Yersinia enterocolitica, Cryptosporidium, Giardia,* or other pathogens or non-infectious agents (11) and is clinically characterized by general abdominal cramping, diarrhea, and nausea (12). In the US, roughly 12-16.4 million cases of AGI attributed to public drinking water systems occur annually (13–15). Although the total burdens of drinking-water associated AGI and other waterborne illnesses in countries with high access to safe drinking water are uncertain (16,17), evidence suggests individuals who use private wells are at exacerbated risk in comparison to those receiving water from public water systems. For example, a study conducted in North Carolina found that, of drinking-water associated AGI emergency department visits, 99% were associated with private well contamination while only 1% were associated with public water systems, reflecting that the burden of drinking-water related AGI falls on private well users (18). During extreme rainfall or flooding, pathogens can be transmitted through agricultural or stormwater runoff or sewer discharge, increasing waterborne disease risk (19–21). Studies globally have documented increases in the incidence of AGI following hurricanes, flooding, and heavy rainfall events (21–24). In NC, prior studies have linked hurricane-associated flooding to increased rates of AGI emergency department visits following Hurricanes Florence and Matthew (20), and increased outpatient visits for waterborne diseases following Hurricane Floyd (25). At the time of this work, studies on the impacts of Hurricane Helene have documented impacts on mental and physical health, infrastructure, communication, and food security in surveys of affected residents (26,27). However, the literature remains limited on adverse health outcomes in NC following Hurricane Helene and which populations were most affected, including residents who use private wells.

This study investigated the impacts of Hurricane Helene on the incidence of AGI and sought to determine whether these impacts were different in areas where more residents use private wells. We used a newly available electronic health record (EHR) database with shorter delays in data availability than traditional government surveillance resources to provide a more expedient understanding of the impacts of the hurricane. We adopted a controlled interrupted time series design for our analyses, allowing us to estimate the effect of Hurricane Helene on the incidence of AGI in flooded regions, controlled for pre-hurricane trends and post-hurricane changes in control regions. Our study provides one of the first analyses of the public health implications of one of the deadliest disasters in NC’s history and furthers the understanding of how extreme events like hurricanes can affect health in rural regions of the US.

## 2. Methods

### 2.1 Study objectives

Our primary study objective was to investigate the impact of Hurricane Helene on the incidence of AGI in NC, US using EHR available from 2017-2024. All analyses were conducted at the zip3 and week (zip3-by-week) level due the spatial granularity available in the EHR data. Here, zip3 units refer to the group comprising five-digit zip codes that share the same first three digits. We also assessed whether impacts of the hurricane were different in areas with greater proportions of residents using private wells. For comparison, we used a similar approach to investigate the impact of Hurricane Florence, which impacted NC in 2018 and has been examined previously using data from the NC Disease Event Tracking and Epidemiologic Collection Tool (NC DETECT) (11,20).

### 2.2 Outcome data

We ascertained cases of AGI in NC at the zip3-by-week level through a data access agreement with Truveta, which provides access to de-identified EHR data on a shorter delay in data availability compared to traditional government data sources. Data from inpatient and outpatient visits were included. Truveta’s data is collected from member healthcare organizations within the US. No data is included from immigration facilities, correctional facilities, military facilities, or Indian health facilities. EHR data for this study were extracted in January 2026 with a time-optimized study profile to provide an appropriate level of temporal accuracy. Truveta’s deidentification is attested to by a qualified expert in accordance with the Health Insurance and Portability Act (HIPAA) Privacy Rule. Truveta’s deidentification process excludes patients ingroups with a high risk of reidentification (e.g., patients with rare diseases, rare ethnic groups). In the time-optimized study profile, this results in data masking into coarser geographic regions, such as at the zip2 or state level. Additional details on Truveta’s data model have been described in detail previously (28,29).

Diagnosis codes from the *International Classification of Diseases, Tenth Revision* (ICD 10) and SNOMED CT were used to classify AGI, following approaches in prior literature on hurricanes, flooding, and AGI hospitalizations (18,20–22). For the ICD 10, these included intestinal infectious diseases (A00-A09), unspecified non-infectious gastroenteritis and colitis (K52.3, K52.89, K52.9), vomiting (R11.10-R11.12), and diarrhea (R19.7). We included equivalent codes from SNOMED CT after consulting with Truveta staff experts (Supplemental Information [SI] Table S1 contains a list), as well as the hierarchical descendants (*e.g.*, children or grandchildren) of all codes. For each zip3, we tabulated the weekly number of AGI visits from 2017-2024 for our analyses. Here, the zip3 locations correspond to where the healthcare interactions for AGI occurred. A total of 547,986 visits for AGI was observed without masking from 2017-2024 at the zip3 level, with an additional 328,834 (60%) that were geographically masked in zip2 or state-level groups.

### 2.3 Flooding exposure datasets

Satellite-derived flood depth estimates were obtained from the Global Flood Monitoring System (GFMS), which estimates flooding extents in three-hour increments at a 1/8^th^ degree resolution (30). Zip code tabulation area (ZCTA) boundaries were obtained from the 2020 U.S. Census and aggregated to the zip3 spatial scale to match the spatial resolution of the outcome data (31). We calculated the zip3 area-weighted mean, median, and maximum percent of flooding over the period of the hurricane to compare inundation metrics (September 24, 2024 12:00AM to September 28, 2024 9:00PM) (6). Prior to conducting the statistical analysis, we defined a zip3 unit as exposed to flooding if, on average, >50% of the area was flooded over three-hour increments throughout the course of the hurricane (hereafter referred to as “exposed” and “control” units). We chose this definition because it allowed for acceptable balance between the exposed and control groups since all zip3 units experienced at least some flooding (see Figure S1). Classification based on the median percent area flooded was also identical.

We calculated the total population in each year for each zip3 unit using the ZCTA-level American Community Survey 5-year estimates for 2017-2023 (32). When this analysis was ongoing, population estimates for 2024 were not yet available, so 2023 population estimates were carried forward for 2024.

Exposure to Hurricane Florence was assessed using the Global Flood Observatory (GFO) (33). The GFO was used because the GFMS did not have data for the time period during which Hurricane Florence took place (September 14-16, 2018) (34). For each flooding event, the GFO dataset includes start and end date, country, area, main cause, number of fatalities, number displaced, a severity score, a flood impact index, and a polygon boundary of the area that experienced flooding. We rasterized the polygon boundary to create a binary raster dataset identifying areas that experienced flooding in North Carolina due to Hurricane Florence (1 = flooded, 0 = not flooded). We then calculated the area in each zip3 that experienced flooding and used the same exposure definition as Hurricane Helene (i.e., >50% of the zip3 unit experiencing flooding).

### 2.4 Covariates

Data on daily temperature (minimum, maximum, and mean), precipitation, and dewpoint were obtained from the Parameter-elevation Regressions on Independent Slopes Model dataset at the 800 meter resolution (35). Covariates were aggregated to the zip3-by-week level, where we calculated the weekly averages of the mean daily temperature, minimum temperature, maximum temperature, precipitation, and the weekly sum of total precipitation. We used dewpoint to calculate the average relative humidity per zip3 unit per week with the Magnus approximation (36). These covariates were used for exploratory data analysis to describe weather patterns in the weeks prior to the hurricane and were additionally considered as potential time-varying confounders in sensitivity analyses.

We used 2020 census-block level domestic water source estimates to obtain the percentage of private well users in each zip3 unit (37). In exploratory analyses, we compared sociodemographic characteristics between the exposed and control zip3 units using data from the American Community Survey 2024 5-year estimates and 2020 decennial US Census (32,38). Variables included racial/ethnic compositional factors, health insurance coverage, number and percent of rural households, median annual income, and land area.

### 2.5 Statistical analyses

All primary analyses used a controlled interrupted time series (CITS) design to estimate the average change in the incidence rates of AGI in exposed zip3 units in the weeks following each hurricane compared to the incidence rate had the hurricane not occurred, controlled for changes in the control areas after the hurricane. This method compares each zip3 to itself over time, which controls for time-invariant zip3 characteristics by design. We fit separate CITS models for each hurricane. We defined three-week, five-week, and eight-week post-hurricane intervals as intercept shifts for both hurricanes. Following prior approaches (20), we removed visits for AGI in the eight-week period following other landfalling hurricanes in NC from the 2017-2024 time series to remove the influence of other major hurricanes in the time series (39,40). For Hurricane Helene, this removed data during Hurricanes Isaiah (in 2020), Dorian (2019), and Florence (2018). Since we used AGI data from 2017-2024 and there were no landfalling hurricanes in NC in 2017, this did not impact the time series for Hurricane Florence.

Our primary models for Hurricanes Helene and Florence were of the following form:

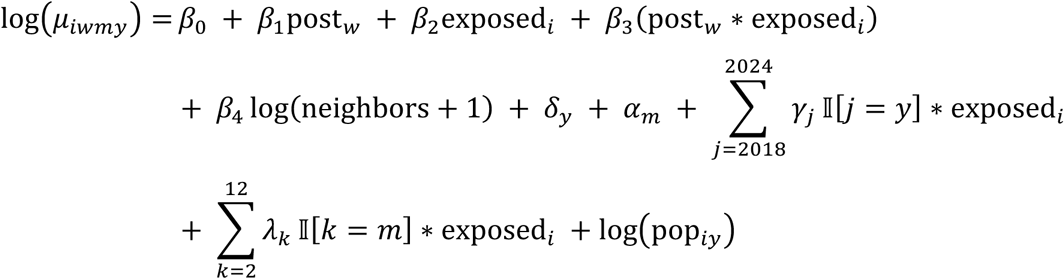

where *μ_iwmy_* is the number of AGI visits in zip3 *i* in week *w* in month *m* and year *y*; *β*_%_ and *β*_’_ are coefficients for the control and the additional difference in the exposed units in the pre-hurricane period, respectively; *β*_&_ and *β*_(_ are coefficients (as three-week, five-week, and eight-week periods after the occurrence of the hurricane) for the control and the additional difference for the exposed zip3 units in the post-hurricane period, respectively; *δ*_$_ and *γ*_*_ are year fixed-effects among the control and the additional yearly difference among exposed units, respectively; *α*_#_ and *λ*_-_ are month fixed-effects among the control and additional monthly difference for exposed units, respectively; and log$pop_!$_& is an offset term for the total population in each zip3 in year *y* from the 5-year ACS estimates. To account for moderate spatial autocorrelation observed in initial models (global Moran’s I ranging from 0.17 to 0.21, *p* = 0.07 to 0.08), we included the number of AGI visits in neighboring zip3 units as a spatially lagged independent variable (shown as log(neighbors + 1)). First order neighbors were identified using queen’s contiguity and neighboring AGI visits were weighted by the number of neighbors for each zip3. In primary models, we accounted for overdispersion in AGI visits by assuming a quasi-Poisson error distribution. Our primary coefficient of interest was *β*_(_, an intercept shift which estimates the change in AGI incidence in exposed compared to control units in the weeks after the hurricane, controlled for pre-hurricane temporal trends.

We examined whether hurricane-associated changes in AGI incidence disproportionately affected areas with residents using private wells in two ways. We examined effect heterogeneity among zip3 units with greater proportions of residents on private wells by introducing product terms between the continuous proportion of private well users in each zip3, the pre-/post-indicator for the occurrence of the hurricane, and the inundation exposure indicator. We also reclassified the CITS model in terms of exposure and control zip3 units according to whether >30% of residents used private wells. We explored other, lower cut-points (such as 10% and 20%) with similar results, but we did not examine an indicator for whether > 50% of residents used private wells because only one zip3 would be classified as such (Figure S2). The SI section 1.1 contains additional methodological details for both sets of analyses.

### 2.6 Examining the impacts of geographic masking

Geographic masking in the EHR database resulted in AGI visits that could not be analyzed directly at the zip3 level and were aggregated into coarser spatial units, such as the zip2 or state level. For our analysis Hurricane Helene, we used a hierarchical allocation procedure to examine the sensitivity of our results to this masking procedure. This approach is comparable to a model-based interpolation of aggregated spatial count data (41), where the allocation model here was fit only to the pre-hurricane time series.

We first generated a new time series with an additional grouping denoting the combination of zip3, week, and compatible, masked geographic regions (zip2 or state level). In our first modeling step, we modeled the incidence of AGI in the pre-hurricane period, accounting for temperature, humidity, and precipitation; year, month, and zip3 fixed-effects; and a variable representing the total number of weeks since the start of the time series. Second, for each combination of zip3, week, and compatible masked geographic unit (zip2- or state-level), we calculated allocation probabilities for the incidence of AGI by dividing the predicted zip3-by-week level visits by the total predicted visits in each compatible masked unit for the full time series. These allocation probabilities were multiplied by the total number of weekly visits for each masked geographic unit to allocate the aggregated visits for each zip3. In our third step, we accounted for uncertainty in the allocation estimates (which can also be fractional) by generating multiply imputed datasets (*n* = 20) where the allocated visits were drawn from a multinomial distribution with parameters obtained from step two (42). This approach preserves the total number of masked AGI visits in each imputed dataset. We then repeated the primary analyses and pooled the estimates and variances across these datasets using Rubin’s rules (43). Additional method details and a model description are provided in the SI section 1.2.

### 2.7 Sensitivity analyses

For our analysis of Hurricane Helene, we conducted several analyses to examine the sensitivity of our results to choices related to model specification, assumptions for the post-hurricane period, and the CITS study design. We compared our primary models that included month fixed-effects to models with season fixed-effects. We separately considered weekly average mean, minimum, and maximum temperature and relative humidity as potential time-varying confounders. Across several years, we observed changes in the AGI visit time series in the October and November periods, which partially co-occurred with the eight-week post-Helene period and may reflect the influence of holidays (Figure S3). We therefore repeated our analysis using a three-week post-hurricane period but excluding the subsequent five weeks from the time series.

As described previously, all zip3 units experienced at least some flooding during Hurricane Helene. We thus repeated our analyses with a non-controlled interrupted time series. These analyses examined whether the occurrence of the hurricane resulted in a significant average increase in the incidence of AGI compared to the incidence rate had the hurricane not occurred. All analyses were conducted in R version 4.5.1 (44).

## 3. Results

We identified 547,986 AGI visits that occurred across 20 zip3 units from 2017-2024. Eight of 20 zip3 units (40%) were classified as exposed to flooding to Hurricane Helene, corresponding to >50% of the land area flooded over the course of the hurricane. Zip3 units that experienced greater flooding during Hurricane Helene comprised approximately 4 million residents (Table S2). These zip3 units comprised regions predominately in western NC and included both rural and urban areas such as Asheville and Wilmington, NC (Figure 1). Compared to control units, exposed zip3 units had greater proportions of non-Hispanic White residents; lower proportions of non-Hispanic Black, Asian, and American Indian and Alaskan Native residents; and higher median household income and population density (Table S2). We observed elevated mean weekly precipitation amongst several exposed and control zip3 units in the weeks preceding and at the start of Hurricane Helene (Figure S4).

**Figure 1:**
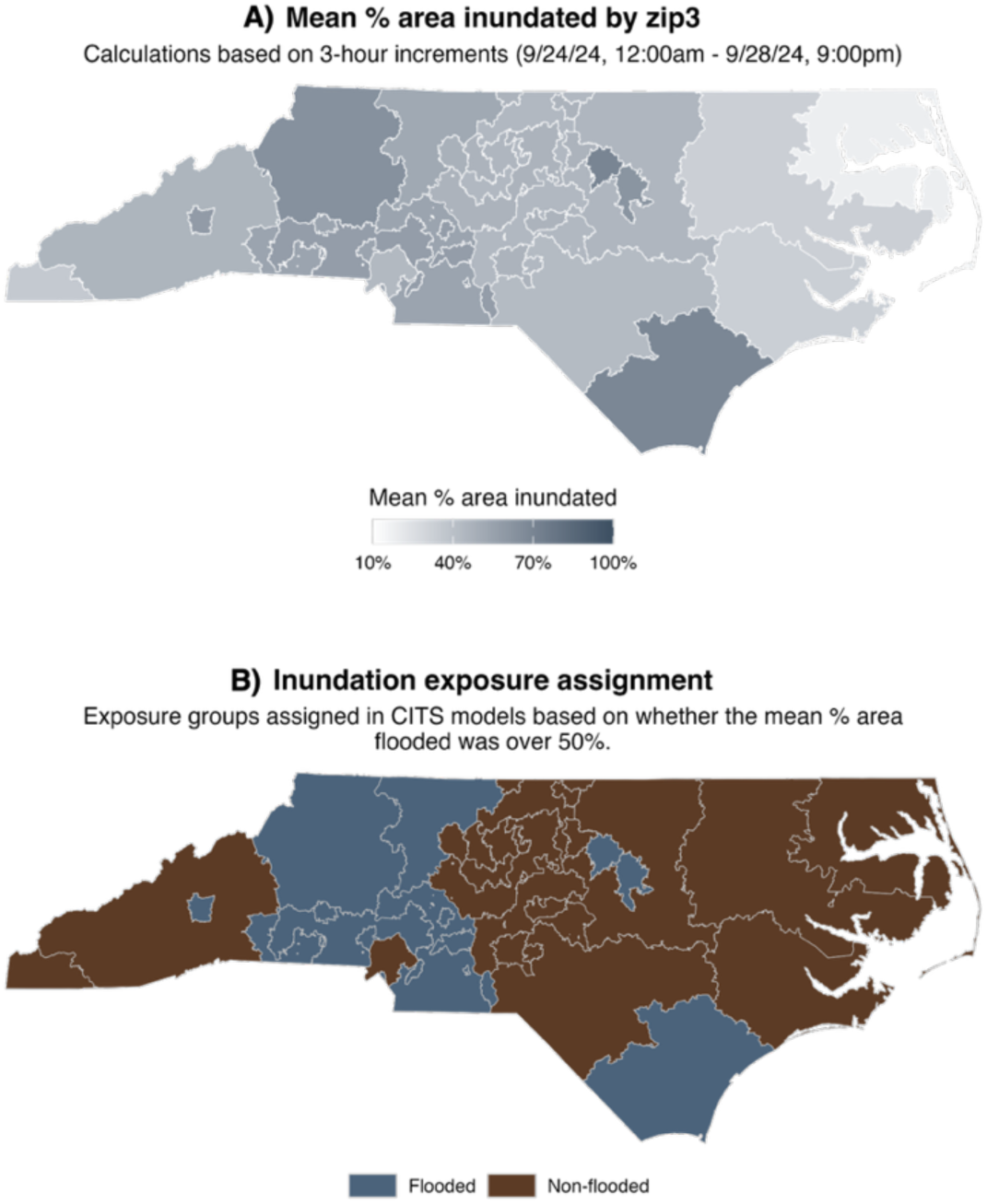
A) Mean percent area inundated for each zip3 over the course of Hurricane Helene calculated from September 24-28, 2024. We used satellite-derived estimates from the Global Flood Monitoring System to quantify inundation during Hurricane Helene (30). B) Hurricane Helene inundation exposure assignment categories at the zip3-level. Exposed (flooded) zip3 units were categorized as such if, on average, a majority of the zip3 was inundated over the course of Hurricane Helene.

### 3.1 Hurricane Helene and the incidence of AGI

Figure 2 shows the time series of AGI visits per 10,000 people in combined exposed and control units from June to December 2024. In the three-week period following Hurricane Helene, we observed an average 11% increase in the incidence of visits for AGI among exposed units in our main model (incidence rate ratio [IRR]: 1.11, 95% confidence interval (CI): 0.59, 2.07) compared to the expected incidence had the hurricane not occurred and controlled for post-hurricane changes among the control units, year and month temporal trends, and spatially lagged AGI visits. However, results were imprecise and non-significant at the 0.05 alpha level. Similar, non-significant increases of 14-16% were observed at the five- and eight-week intervals following the hurricane (Table 1).

**Figure 2:**
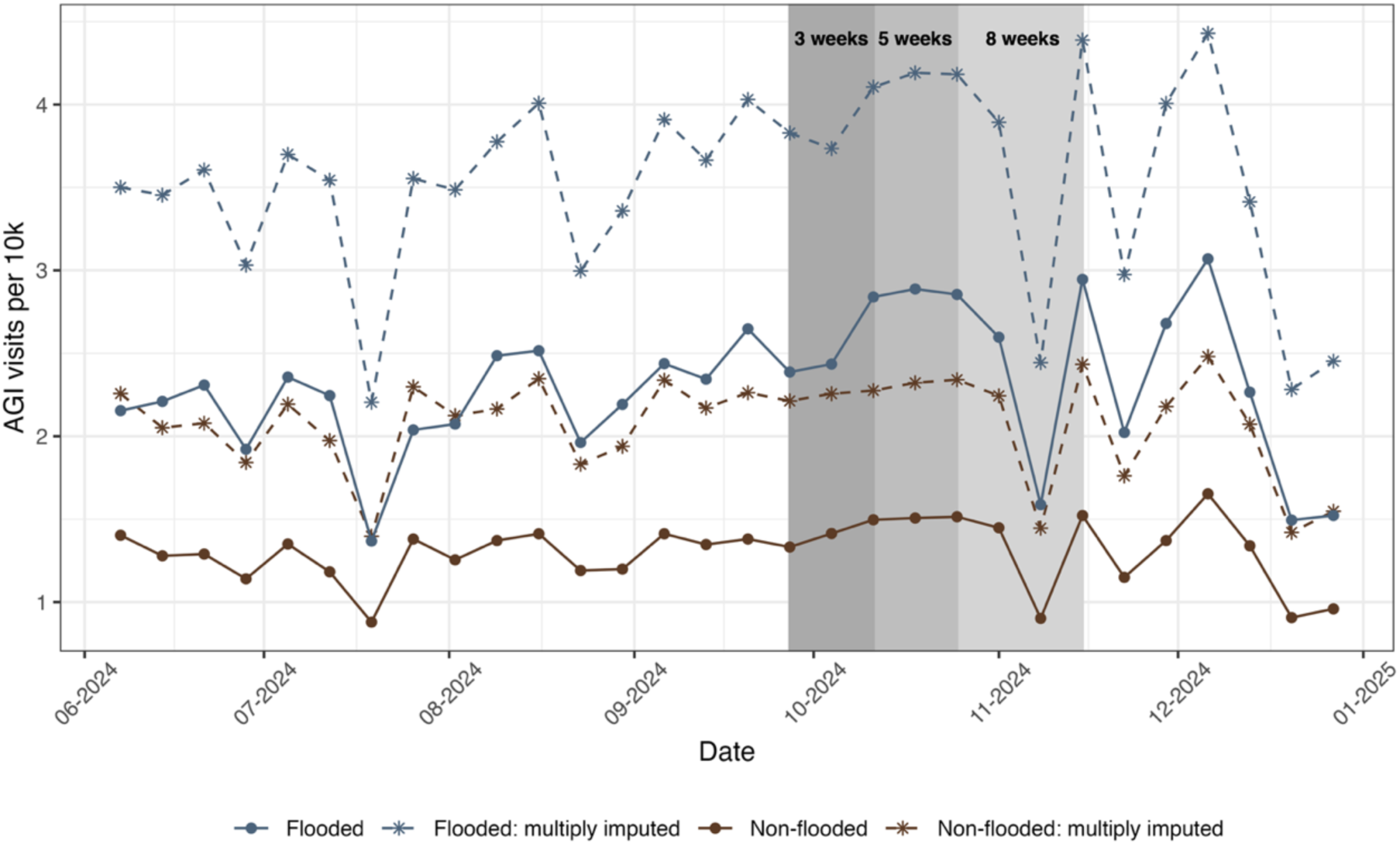
Time series of visits for acute gastrointestinal illness (AGI) in flooded (exposed) and non-flooded (control) zip3 units prior and during Hurricane Helene. Solid lines depict the total visits for AGI divided by the total populations amongst all exposed and control units, respectively. Dotted lines depict the time series accounting for geographically masked visits, where the median total visits across 20 imputed datasets are shown for the exposed and controls units divided by their total populations, respectively. The three-, five-, and eight-week periods following Hurricane Helene are shown with grey shading.

**Table 1:** Incidence rate ratios and 95% confidence intervals for acute gastrointestinal illness (AGI) visits in North Carolina zip3 units following Hurricane Helene (September 2024)

| **Analysis** | **Interval after Hurricane Helene** | **Incidence rate ratio** | **Total AGI visits** | **Sample size** |
|---|---|---|---|---|
| Primary models<sup>a</sup> | Three-week | 1.11 [0.59, 2.07] | 547,986 | 7,940 |
|  | Five-week | 1.14 [0.69, 1.91] |  |  |
|  | Eight-week | 1.16 [0.76, 1.76] |  |  |
| Models with imputed datasets<sup>b</sup> | Three-week | 1.07 [0.56, 2.03] | 876,820 |  |
|  | Five-week | 1.09 [0.64, 1.86] |  |  |
|  | Eight-week | 1.10 [0.71, 1.70] |  |  |

Results in this table correspond to exp(*β*<sub>3</sub>) from the primary models evaluating different intervals after the start of the hurricane. Results are from quasi-Poisson controlled interrupted time series (CITS) models at the zip3-by-week level that included an offset for total population and adjustment for year and month fixed-effects and a spatially lagged based on AGI visits in neighboring units. Zip3 units were defined as exposed if, on average, a majority of their area was flooded over the course of Hurricane Helene. The sample size refers to the total zip3-by-week observations.
<sup>a</sup>These refer to results from the main CITS models that do not consider geographically masked AGI visits in the underlying electronic health record dataset.
<sup>b</sup>These refer to pooled results from models fit to 20 imputed datasets that incorporated geographically masked AGI visits. AGI visits were spatially disaggregated into zip3 units using a hierarchical allocation procedure.

The time series for the multiply imputed datasets accounting for geographically masked AGI visits were generally parallel in temporal trends in comparison to the main analyses that did not account for masking (Figure 2). We also observed similar IRRs after pooling results across CITS models that accounted for geographically masked AGI visits (Table 1). For example, in the three-week interval following Hurricane Helene, the IRR for AGI among exposed zip3 units was 1.10 (95% CI: 0.56, 2.03).

We observed similar results in analyses examining sensitivity of the primary CITS models to differences in model specification and the post-Helene period (Table S3). For example, we observed 9-16% increases in the incidence of visits for AGI among exposed units in negative binomial models. Confidence intervals in these sensitivity analyses were similarly imprecise and included the null. In non-controlled ITS analyses, effect estimates were attenuated in comparison to the CITS models with confidence intervals including the null (*e.g*., IRR in the three-week interval: 1.00 95% CI [0.71, 1.37]; Table S4).

### 3.2 Analyses examining impacts on residents using private wells

Table S5 shows coefficient estimates from models including additional product terms to examine effect heterogeneity related to private well use and models with a redefined exposure classification based on the proportion of residents using private wells. We did not observe significant increases in the incidence of AGI among zip3 units with >30% residents using private wells compared to control units in the weeks following Hurricane Helene (Table S5). For example, in the three weeks following the hurricane, we observed an 8% decrease in the incidence of AGI visits among zip3 units with >30% residents using private wells (IRR: 0.92, 95% CI: 0.35, 2.10), compared to pre-hurricane trends and controlled for post-hurricane changes in the control units. We also generally did not observe evidence of significant heterogeneity in the impact of Hurricane Helene on flooding units in association with the proportion of residents using private wells (Table S4). One exception was for the eight-week interval following the hurricane (IRR for the three-way product term: 0.98 [0.95, 1.00], *p* = 0.07). However, in this model, we did not observe significant shifts in AGI visits following the hurricane among any of the additional two-way product terms.

### 3.3 Hurricane Florence and the incidence of AGI

Table S6 shows coefficient estimates from models examining the impact of Hurricane Florence in September 2018 on the incidence of AGI visits in zip3 units exposed to hurricane-related flooding. Effect estimates here indicated increases of 6-11% among the exposed zip3 units at the three-, five-, and eight-week intervals following the hurricane. However, results were similarly imprecise, with wide 95% confidence intervals that included the null.

## 4. Discussion

Our study investigated the impacts of Hurricane Helene on the incidence of visits for AGI in NC at the zip3 and week level. Using a CITS study design, we observed 11-16% increases in the incidence for AGI visits among more heavily flooded zip3 units after the hurricane, albeit estimates were imprecise with wide confidence intervals including the null. We did not observe clear evidence that the hurricane disproportionately impacted areas with more residents using private wells. Results were consistent across several sensitivity analyses, and similar results were observed for Hurricane Florence, which made landfall in NC in 2018.

Prior literature on weather-related impacts on the incidence of AGI globally have primarily focused on heavy rainfall, temperature, and flooding, although fewer have focused specifically on hurricane-associated flooding (23). Among studies focused on heavy rainfall events, several have reported significant increases and outbreaks in diarrheal and waterborne diseases, including nationwide and regional studies situated in the US (45), United Kingdom (46), Canada (19,47), Eswatini (48), Japan (49), and China (50). Prior reviews have found that a commonality among these outbreaks is the occurrence of heavy rainfall preceded by a dry period, a phenomenon in line with the “concentration-dilution hypothesis” (23,24). Under this hypothesis, pathogens can accumulate during periods of less rain, while heavy rainfall can flush pathogens into surface water bodies. However, longer periods of heavy rainfall may dilute pathogen concentrations, reducing risk of waterborne diseases. In our study, we observed elevated rainfall prior to Hurricane Helene in several zip3 units (Figure S4), coinciding with prior storms earlier in September 2024 (51), which may partially explain our findings.

Studies on the impacts of prior hurricanes on AGI incidence in NC have typically relied on data from the North Carolina Disease Event Tracking and Epidemiologic Collection Tool (NC DETECT) (20) or Medicaid claims (25). For example, Quist *et al*., 2022 employed a similar CITS study design and observed an 11% increase in AGI emergency department visits after Hurricanes Matthew and Florence (20). A follow-up 2025 study examined the differential impacts of hurricane-associated flooding in areas with more concentrated animal feeding operations (CAFOs), finding increases AGI emergency department visits in zip5 units with more CAFOs following Hurricane Florence (11). In NC, CAFOs follow distinct geographical and social patterning, with more hog CAFOs in rural and eastern counties with higher proportions of people of color, concentrated poverty, and multiple other environmental hazards (*e.g*., landfills) (20,52,53).

Previous studies documenting significant increases in AGI following hurricanes in NC are supported by evidence that these storms result in elevated concentrations of various pathogens and contaminants in surface water bodies (54,55) and microbial contamination of drinking water sources, particularly private wells (4,10). At the time of this work, studies on Hurricane Helene are scarce. However, a mixed-method survey indicated that residents in Southern Appalachia most frequently reported health conditions that suggested exposure to contaminated water and mold, including headaches; nose, eye, skin, and throat irritation or cough; and gastrointestinal issues (27). Voluntary well testing following Hurricane Helene revealed that approximately 30% of private wells in western counties of NC tested positive for total coliform (10,56), although these testing programs did not detail specific pathogen species. Aside from exposures to contaminated water and related health conditions, surveys of affected residents have documented widespread impacts on mental health, physical infrastructure, and food security following the hurricane (26,27).

Our study was strengthened in its use of an EHR data source with a relatively short lag time, allowing us to study Hurricane Helene on a quicker timeline in comparison to traditional government data sources. We employed a quasi-experimental study design, which reduces confounding by time-invariant factors. The CITS framework further accounts for potential common shocks coincident with the hurricane that may have impacted both exposed and units. Satellite-derived estimates were used to classify exposure groups for two hurricanes according to similar definitions. Additionally, we were able to examine almost 8 years of pre-Hurricane Helene AGI trends.

However, our study faced several limitations. Many individuals affected by AGI in rural communities may not seek health care resulting in undercounting and potential under-estimation of the effect of hurricanes on AGI. During extreme weather events, individuals across the state may also change healthcare seeking behaviors or experience disruptions in care.(57) Although state coverage for Truveta in NC is high (>100% of the census population accounted for across encounters in NC), the data are limited in both spatial and temporal granularity. Prior studies on earlier hurricanes have reported similar effect sizes to those observed here that were significant at the 0.05 level, although with data available at finer spatial and temporal scales such as the zip5-by-day level (20). Truveta’s database also includes only data from participating member health systems, which may not be representative of all parts of the state. While we attempted to account for geographic data masking in the AGI visit data obtained from Truveta, we cannot account for unmeasured factors that contributed to masking, which may be a particular concern for lower counts observed in lesser populated areas. Finally, we were unable to account for population displacement or public health interventions (*e.g.*, issuance of boil water notices) that occurred during and following the hurricane (58) and may have reduced exposure to contaminated water. **Conclusions**

We observed positive, statistically insignificant increases in the incidence of AGI visits in the three- to eight-week window following Hurricane Helene. We used a novel EHR data source with a shorter lag time, although limitations in its spatial and temporal resolution suggest that future work focused on hurricane-associated AGI should examine these relationships at finer scales. Although we did not observe clear evidence indicating that regions with greater proportions of residents using private wells were disproportionately impacted, future work should re-examine these relationships for confirmation because individuals using private wells may be uniquely susceptible to water quality contamination following hurricanes. Additionally, future work should examine the impact of public health interventions employed during hurricanes and whether this may mediate or modify observed relationships. Within quasi-experimental frameworks, comparisons of exposure classifications at different spatial scales would be valuable, particularly within rural areas where private wells are commonly used and may experience highly localized flooding-induced contamination. In the eastern US, the intensity and frequency of hurricanes are expected to increase as the climate changes (59). Thus, studies should continue to examine hurricane-associated water quality contamination and other health outcomes with robust study designs to examine both pre- and post-hurricane data.

## Author contributions

**JML:** Methodology, Formal analysis, Investigation, Data curation, Visualization, Writing - Original Draft, Writing - Review & Editing. **KRO:** Formal analysis, Investigation, Data curation, Visualization, Writing - Original Draft, Writing - Review & Editing. **AP:** Writing - Original Draft, Writing - Review & Editing. **AI:** Writing - Original Draft, Writing - Review & Editing. **TA:** Methodology, Writing - Review & Editing, Funding acquisition. **PB:** Methodology, Writing - Review & Editing. **AP:** Data curation, Writing - Review & Editing. **XCH:** Conceptualization, Methodology, Writing - Review & Editing, Supervision, Funding acquisition, Supervision.

## Data Availability

Aggregated zip3-by-week analytic datasets will be made available after peer review. All R scripts for data processing, analysis, and table/figure generation are available here: https://github.com/water-health-opportunity-lab/hurricane-AGI

## Acknowledgments

This project was supported by a pilot grant from the Research and Engagement for Action on Climate and Health (REACH) Center, funded by National Institute of Environmental Health Sciences (P20 ES036775).

