## Supplementary material for "The Impact of Hurricane Helene on the Incidence of Acute Gastrointestinal Illness in North Carolina, US: A Controlled Interrupted Time Series Analysis": SI

### Table of contents

*Figure and table titles below are linked to their locations in this document.*

*Supplemental methods*
Analyses examining impacts on residents using private wells pg. S3-S4
Hierarchical allocation procedure to address geographic masking limitations pg. S4-S5
*Supplemental tables*
Table S 1: SNOMED CT codes used to ascertain visits for acute gastrointestinal illness (AGI) over 2017-2024 in North Carolina within the Truveta electronic health records database pg. S6
Table S 2: Sociodemographic summary statistics for exposed and unexposed zip3 regions pg. S7
Table S 3: Results from sensitivity analyses (incidence rate ratios and 95% confidence intervals) examining the incidence of visits for acute gastrointestinal illness (AGI) visits in North Carolina zip3 units following Hurricane Helene (September 2024) pg. S8
Table S 4: Results from non-controlled interrupted time series analyses examining changes in the incidence of visits for acute gastrointestinal illness (AGI) in North Carolina zip3 units following Hurricane Helene (September 2024) pg. S9
Table S 5: Results from analyses examining effect heterogeneity related to the proportion residents using private wells for the impact of Hurricane Helene on visits for acute gastrointestinal illness (AGI) in North Carolina zip3 units (September 2024) pg. S10
Table S 6: Results from analyses (incidence rate ratios and 95% confidence intervals) examining changes in the incidence of visits for acute gastrointestinal illness (AGI) in North Carolina zip3 units following Hurricane Florence (September 2018) pg. S11
*Supplemental figures*
Figure S 1: Comparison of inundation exposure metrics across zip3 units (n = 20) during Hurricane Helene pg. S12
Figure S 2: Map and histogram of the estimated proportion of residents using private wells across zip3 units in North Carolina pg. S13
Figure S 3: Yearly time series of total visits for acute gastrointestinal illness ([AGI], per 10k people) across North Carolina (NC) pg. S14
Figure S 4: Daily time series of mean precipitation among exposed and control units from September to December 2024 pg. S15
Figure S 5: Zip3 weekly trends in AGI visits (per 10k) from June to December 2024 pg. S16
Figure S 6: Zip3-by-week level trends in AGI visits (per 10k) in the imputed datasets (m = 20) from June to December 2024 pg. S17
*References* pg. S18

### *1. Supplemental methods*

#### 1.1. Analyses examining impacts on residents using private wells

*Model 1: Effect heterogeneity of Hurricane Helene flooding related to private well use*

As described in the main manuscript, this model retains the exposure classification from the primary models but adds additional product terms with a variable pertaining to the proportion of private wells users in each zip3 ($\text{prop\_well}_i$ below). Additional model details are available in the main manuscript.

$$\begin{aligned} \log(\mu_{iwmy}) = {} & \beta_0 + \beta_1 \text{post}_w + \beta_2 \text{exposed}_i + \beta_3 \text{prop\_well}_i + \beta_4 (\text{post}_w * \text{exposed}_i) \\ & + \beta_5 (\text{exposed}_i * \text{prop\_well}_i) + \beta_6 (\text{post}_w * \text{prop\_well}_i) \\ & + \beta_7 (\text{post}_w * \text{exposed}_i * \text{prop\_well}_i) + \beta_8 \log(\text{neighbors} + 1) + \delta_y + \alpha_m \\ & + \sum_{j=2018}^{2024} \gamma_j \, \mathbb{I}[j = y] * \text{exposed}_\text{i} + \sum_{k=2}^{12} \lambda_k \, \mathbb{I}[k = m] * \text{exposed}_\text{i} \\ & + \log(\text{pop}_{iy}) \end{aligned}$$

In this model:

- $\mu_{iwmy}$ is the number of AGI visits in zip3 *i* in week *w* in month *m* and year *y*
- Among zip3 units without private well use, $\beta_0$ and $\beta_2$ are coefficients for the pre-hurricane period for the control and the additional difference for exposed units, respectively
- Among zip3 units without private well use, $\beta_1$ is the coefficient for the post-hurricane period among the control units
- $\beta_3$ is the incremental change in AGI incidence for the proportion of residents using private wells in the pre-hurricane period among the control units
- Among zip3 units without private well use, $\beta_4$ is the additional shift in AGI incidence among exposed units in the post-hurricane period
- $\beta_5$ is the additional incremental change in AGI incidence for the proportion of residents using private wells in the pre-hurricane period among the exposed units
- $\beta_6$ is the incremental change in AGI incidence for the proportion of residents using private wells in the post-hurricane period among the control units
- $\beta_7$ is the additional incremental change in AGI incidence for the proportion of residents using private wells in the post-hurricane period among the exposed units
- $\beta_8$ is a spatially lagged variable based on first-order neighbors with a queen's contiguity matrix
- $\delta_y$ and $\gamma_j$ are year fixed-effects among the control and the additional yearly difference among exposed units, respectively
- $\alpha_m$ and $\lambda_k$ are month fixed-effects among the control and additional monthly difference for exposed units, respectively
- $\log(\text{pop}_{iy})$is an offset term for the total population in each zip3 in year *y* from the 5-year ACS estimates.

*Model 2: Alternate controlled interrupted time series models with redefined exposure groups*

This model redefines the exposure groups from the primary models and Model 1 above with a new exposure variable ($\text{high\_wells}_i$) corresponding to whether >30% of the residents in the zip3 use private wells.

$$\log(\mu_{iwmy}) = \beta_0 + \beta_1 \text{post}_w + \beta_2 \text{high\_wells}_i + \beta_3 (\text{post}_w * \text{high\_wells}_i) + \beta_4 \log(\text{neighbors} + 1) + \delta_y + \alpha_m + \sum_{j=2018}^{2024} \gamma_j \, \mathbb{I}[j = y] * \text{exposed}_\text{i} + \sum_{k=2}^{12} \lambda_k \, \mathbb{I}[k = m] * \text{exposed}_\text{i} + \log(\text{pop}_{iy})$$

In this model:

- $\mu_{iwmy}$ is the number of AGI visits in zip3 *i* in week *w* in month *m* and year *y*
- $\beta_0$ and $\beta_2$ are coefficients for the pre-hurricane period for the control and exposed units (classified as having greater than >30% residents using private wells), respectively
- $\beta_1$ and $\beta_3$ are coefficients for the post-hurricane period among the control and exposed zip3 units, respectively
- $\beta_4$ is a spatially lagged variable based on first-order neighbors with a queen's contiguity matrix
- $\delta_y$ and $\gamma_j$ are year fixed-effects among the control and the additional yearly difference among exposed units, respectively
- $\alpha_m$ and $\lambda_k$ are month fixed-effects among the control and additional monthly difference for exposed units, respectively
- and $\log(\text{pop}_{iy})$ is an offset term for the total population in each zip3 in year *y* from the 5-year ACS estimates.

### 1.2. Hierarchical allocation procedure to address geographic masking limitations

To address sensitivity to geographic masking in the electronic health records (see the main manuscript, Section 2.6), we used an imputation procedure to model the relative incidences of acute gastrointestinal illness (AGI) at the zip3-by-week level to disaggregated masked cases at either the zip2- or state level. To do so, we first used all pre-Hurricane Helene data to fit the following quasi-Poisson regression model:

$$\log(\mu_{iwmy}) = \beta_0 + \beta_1 \text{week}_{wmy} + \beta_2 \text{temp}_{iwmy} + \beta_3 \text{humidity}_{iwmy} + \beta_4 \text{precip}_{iwmy} + \log(\text{pop}_{iy}) + \delta_y + \alpha_m + \zeta_i$$

where $\mu_{iwmy}$ refers to the zip3-by-week level cases; $\beta_1 \text{week}_{wmy}$ refers to the number of total weeks since the beginning of study period and its coefficient; $\beta_2$temp, $\beta_3 \text{humidity}_{iwmy}$, and $\beta_4 \text{precip}_{iwmy}$ refer to average temperature, humidity, and precipitation and their coefficients, respectively; $\log(\text{pop}_{iy})$ is an offset term for the log of the total zip3-level population; and $\delta_y$, $\alpha_m$, and $\zeta_i$ refer to fixed-effects for year, month, and zip3, respectively.

For the full time series, we then estimated the relative incidence of AGI cases as allocation probabilities using the predicted zip3-by-week level cases from the first step:

$$p_{iwmy} = \frac{\hat{\mu}_{iwmy}}{\sum_{k=1}^{K} \hat{\mu}_{kwmy}}$$

where $k = 1, 2, \ldots, K$ denotes the zip3 units compatible with each zip2- or state-level total. For example, the 280 through 289 zip3 units are compatible with 28 (a zip2 unit). For each combination of zip3, week, and compatible masked geographic unit, we then multiplied the relative incidence for each zip3-by-week unit by the total masked cases to disaggregate the masked cases across the compatible zip3s.

To account for uncertainty in the imputed estimates (which can also be fractional), we created multiple ($n = 20$) imputed datasets. Each zip3's weekly imputed estimate was obtained using draws from a multinomial distribution where the probabilities were set to $p_{iwmy}$ and the number of trials was set to $\sum_{k=1}^{K} \hat{\mu}_{kwmy}$ (each set of masked compatible AGI weekly cases). Per the main manuscript, we then repeated the primary analyses for Hurricane Helene and pooled the estimates and variances across these datasets using Rubin's rules.

### 2. *Supplemental tables*

**Table S 1:** SNOMED CT codes used to ascertain visits for acute gastrointestinal illness (AGI) over 2017-2024 in North Carolina within the Truveta electronic health records database

| SNOMED CT code | Condition name |
|---|---|
| 63650001 | Cholera |
| 154269008 | Cholera |
| 4834000 | Typhoid fever |
| 302231008 | Salmonella infection |
| 36188001 | Shigellosis |
| 66107000 | Bacterial food poisoning |
| 154273006 | Bacterial food poisoning |
| 111910009 | Amebic infection |
| 58265007 | Giardiasis |
| 240370009 | Cryptosporidiosis |
| 721810002 | Infection caused by *Cyclospora* |
| 89933001 | Protozoal intestinal disease |
| 186123006 | Other protozoal intestinal diseases |
| 371423007 | Isosporiasis |
| 57725006 | Balantidiasis |
| 56722001 | Infection by *Balantidium coli* |
| 78420004 | Viral enteritis of intestine |
| 186156007 | Infectious colitis, enteritis, and gastroenteritis |
| 235746007 | Indeterminate colitis |
| 309755003 | Other specified non-infective gastroenteritis and colitis |
| 43240000 | Diarrhea of presumed infectious origin |
| 146291000119108 | Vomiting without nausea |
| 8579004 | Projectile vomiting |
| 139338000 | Projectile vomiting |

We also included all hierarchical descendants of the codes above to capture a broader group of visits for AGI. The same procedure was applied for the ICD10 codes listed in the main manuscript.

**Table S 2:** Sociodemographic summary statistics for exposed and unexposed zip3 regions

| | Exposed to flooding (*n* = 8) | Not exposed to flooding (*n* = 12) |
|---|---|---|
| Total population | 4,014,500 | 6,716,000 |
| Area [sqmi] | 15,240 | 34,350 |
| Population density [people per sqmi] | 263 | 196 |
| **Racial and ethnic composition** | | |
| Non-Hispanic Black | 581,900 (14.5%) | 1,564,800 (23.3%) |
| Non-Hispanic White | 2,724,100 (67.9%) | 3,714,600 (55.3%) |
| Non-Hispanic Asian | 115,500 (2.9%) | 235,500 (3.5%) |
| Non-Hispanic American Indian or Alaskan Native | 7,500 (0.2%) | 79,100 (1.2%) |
| Non-Hispanic two or more races | 159,000 (4.0%) | 275,900 (4.1%) |
| Hispanic/Latino | 407,700 (10.2%) | 806,100 (12%) |
| **Other sociodemographic factors** | | |
| Median household income [USD] | 78,000 ± 10,300 | 69,300 ± 12,800 |
| Number of rural households | 560,100 (31.8%) | 1,053,500 (35.7%) |
| No health insurance | 371,000 (9.3%) | 665,500 (10.2%) |

Data were obtained from the 2024 American Community Survey (ACS) 5-year estimates,[1] with the exception of number of rural households, which was obtained from the 2020 decennial US Census Demographic and Housing Characteristics file.[2] Number of rural households refers to the number of households designated as rural by the US Census Bureau. No health insurance is the number and percent of individuals without health insurance.

**Table S 3:** Results from sensitivity analyses (incidence rate ratios and 95% confidence intervals) examining the incidence of visits for acute gastrointestinal illness (AGI) visits in North Carolina zip3 units following Hurricane Helene (September 2024)

| **Sensitivity analysis** | **Interval after Hurricane Helene** | **Incidence rate ratio** | **Total AGI visits** | **Sample size** |
|---|---|---|---|---|
| Model with season fixed-effects | Three-week | 1.13 [0.62, 2.07] | 547,986 | 7940 |
| Negative binomial models | Three-week<br>Five-week<br>Eight-week | 1.09 [0.39, 3.23]<br>1.15 [0.49, 2.75]<br>1.16 [0.59, 2.35] | 547,986 | 7940 |
| Add mean daily temperature and humidity | Three-week | 1.09 [0.58, 2.04] | 547,986 | 7940 |
| Add mean daily temperature and humidity | Three-week | 1.09 [0.58, 2.05] | 547,986 | 7940 |
| Add mean daily temperature and humidity | Three-week | 1.09 [0.58, 2.04] | 547,986 | 7940 |
| Excluding five-week period after initial three weeks | Three-week | 1.14 [0.61, 2.15] | 538,325 | 7840 |

Results in this table correspond to exp($\beta_3$), similar to the primary model description in the main manuscript. Results are from quasi-Poisson controlled interrupted time series (CITS) models at the zip3-by-week level that included an offset for total population and adjustment for year and month fixed-effects and a spatially lagged based on AGI visits in neighboring units. Zip3 units were defined as exposed if, on average, >50% of their area was flooded over the course of Hurricane Helene. The sample size refers to the total zip3-by-week observations.

**Table S 4:** Results from non-controlled interrupted time series analyses examining changes in the incidence of visits for acute gastrointestinal illness (AGI) in North Carolina zip3 units following Hurricane Helene (September 2024)

| **Analysis** | **Interval after Hurricane Helene** | **Incidence rate ratio** | **Total AGI visits** | **Sample size** |
|---|---|---|---|---|
| Non-controlled interrupted time series models | Three-week | 1.00 [0.71, 1.37] | 547,986 | 7,940 |
| | Five-week | 1.03 [0.78, 1.34] | | |
| | Eight-week | 1.02 [0.81, 1.27] | | |

Results in this table correspond to the coefficient describing the post-Hurricane Helene period among all zip3 units. Results are from quasi-Poisson non-controlled interrupted time series (CITS) models at the zip3-by-week level that included an offset for total population and adjustment for year and month fixed-effects and a spatially lagged based on AGI visits in neighboring units. The sample size refers to the total zip3-by-week observations.

**Table S 5:** Results from analyses examining effect heterogeneity related to the proportion residents using private wells for the impact of Hurricane Helene on visits for acute gastrointestinal illness (AGI) in North Carolina zip3 units (September 2024)

| **Model** | **Interval after Hurricane Helene** | **Regression coefficient** | **Incidence rate ratio** |
|---|---|---|---|
| Model 1[a] | Three-week | $\beta_1$ | 0.90 [0.61, 1.29] |
| | | $\beta_4$ | 1.50 [0.67, 3.19] |
| | | $\beta_6$ | 1.01 [0.98, 1.04] |
| | | $\beta_7$ | 0.98 [0.94, 1.02] |
| | Five-week | $\beta_1$ | 0.91 [0.67, 1.23] |
| | | $\beta_4$ | 1.52 [0.82, 2.76] |
| | | $\beta_6$ | 1.01 [0.99, 1.04] |
| | | $\beta_7$ | 0.98 [0.95, 1.01] |
| | Eight-week | $\beta_1$ | 0.90 [0.70, 1.14] |
| | | $\beta_4$ | 1.55 [0.94, 2.52] |
| | | $\beta_6$ | 1.01 [0.99, 1.03] |
| | | $\beta_7$ | 0.98 [0.95, 1.00] |
| Model 2[b] | Three-week | $\beta_3$ | 0.92 [0.35, 2.10] |
| | Five-week | | 0.97 [0.46, 1.93] |
| | Eight-week | | 0.96 [0.53, 1.70] |

Refer to SI section 1.1 for a description of each regression coefficient. Results for each coefficient are exponentiated in the table above.

[a]This pertains to a model that included product terms to examine effect heterogeneity of the impact of Hurricane Helene on exposed zip3 units with greater proportions of private well users.

[b]This pertains to a CITS model in which the flooding exposure and control groups from the main analysis were replaced with an indicator pertaining to whether >30% of residents used private wells based on estimates from Murray *et al.*, 2025.[3]

**Table S 6:** Results from analyses (incidence rate ratios and 95% confidence intervals) examining changes in the incidence of visits for acute gastrointestinal illness (AGI) in North Carolina zip3 units following Hurricane Florence (September 2018)

| Analysis | Interval after Hurricane Helene | Incidence rate ratio | Total AGI visits | Sample size |
|---|---|---|---|---|
| Models examining Hurricane Florence | Three-week | 1.09 [0.35, 3.53] | 113,362 | 2120 |
| | Five-week | 1.06 [0.46, 2.54] | | |
| | Eight-week | 1.11 [0.53, 2.38] | | |

Results in this table correspond to $\exp(\beta_3)$, similar to the primary model description in the main manuscript. Results are from quasi-Poisson controlled interrupted time series (CITS) models at the zip3-by-week level that included an offset for total population and adjustment for year and month fixed-effects and a spatially lagged based on AGI visits in neighboring units. Zip3 units were defined as exposed if, on average, a majority of their area was flooded over the course of Hurricane Florence. The sample size refers to the total zip3-by-week observations.

### *3. Supplemental figures*

**Figure S 1:** Comparison of inundation exposure metrics across zip3 units (*n* = 20) during Hurricane Helene (September 2024)

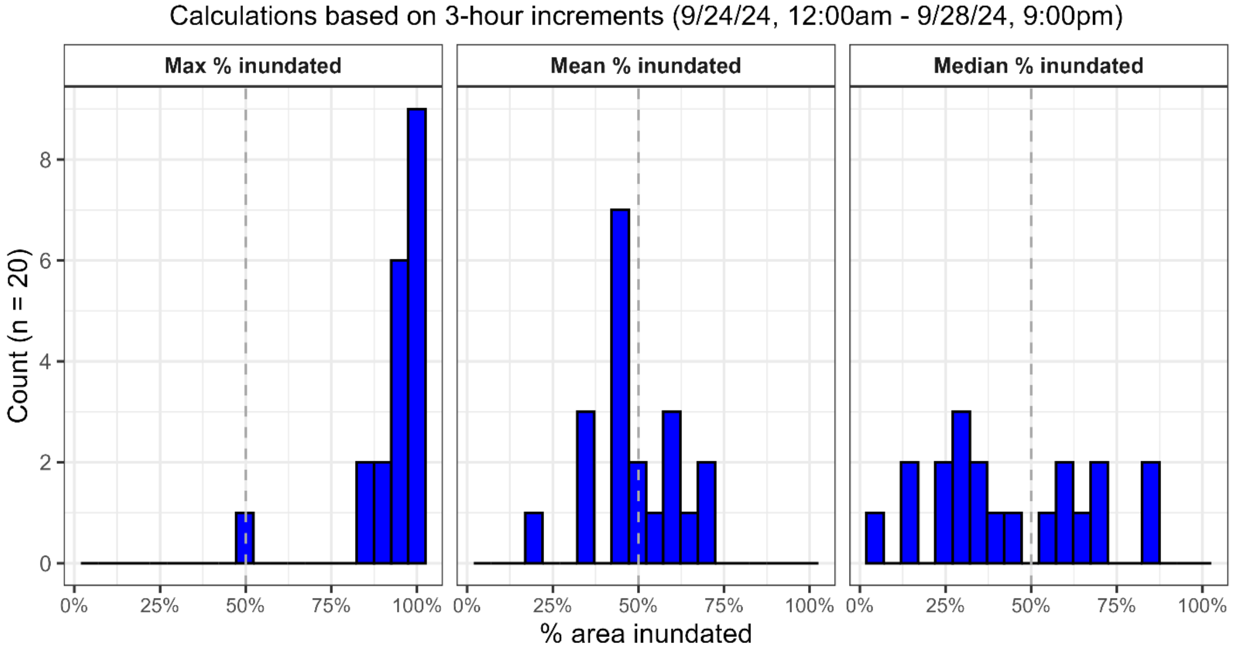


We used satellite-derived estimates from the Global Flood Monitoring System to quantify inundation during Hurricane Helene for zip3 units (*n* = 20) in North Carolina.[4] This figure shows the maximum, mean, and median percent zip3 area inundated over the course of the hurricane based on three-hour increments.

**Figure S 2:** Map and histogram of the estimated proportion of residents using private wells across zip3 units in North Carolina (NC)

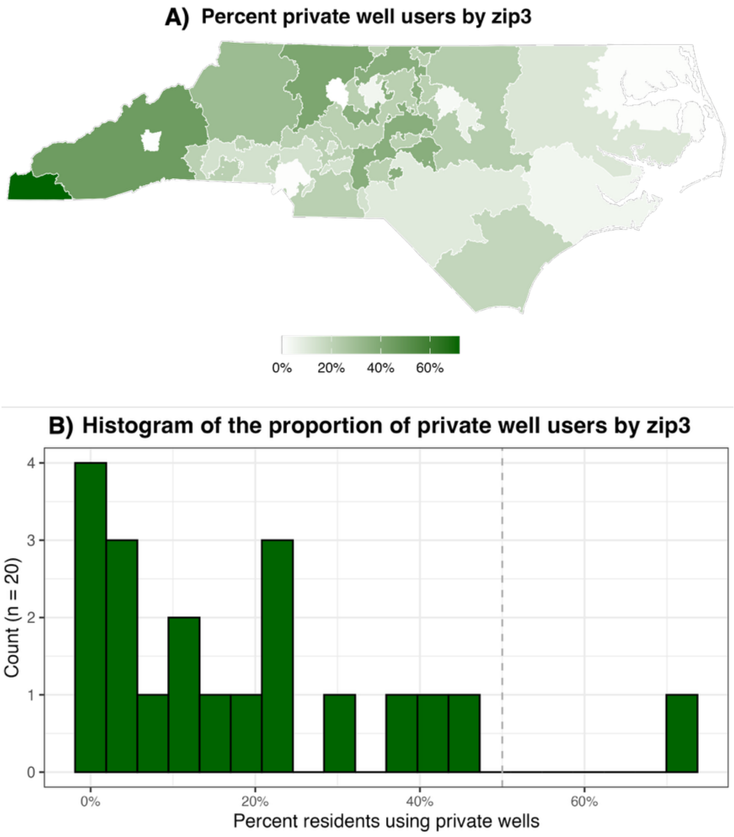


A) Proportion of residents using private wells in each zip3 ($n$ = 20) in NC. The proportion of residents using private wells was calculated using census block estimates from Murray *et al.*, 2025.[3] B) Histogram of the proportion of residents using private wells in each zip3 ($n$ = 20). A dashed line is shown at 50%.

**Figure S 3:** Yearly time series of total visits for acute gastrointestinal illness ([AGI], per 10k people) across North Carolina (NC)

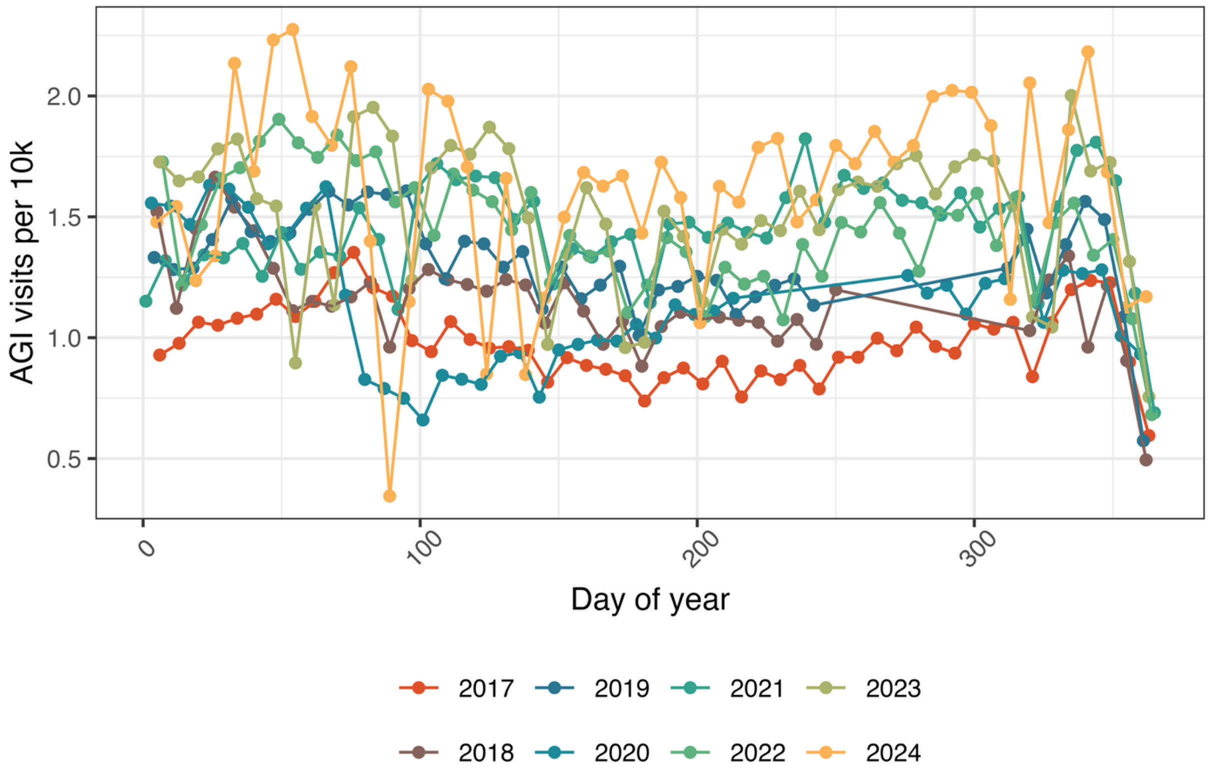


Electronic health records data were obtained through a data access agreement with Truveta. As described in the main manuscript, we removed visits for AGI in the eight-week period following other landfalling hurricanes in NC from the 2017-2024 time series to remove the influence of other major hurricanes in the time series when examining impacts of Hurricane Helene.

**Figure S 4:** Daily time series of mean precipitation among exposed and control units from September to December 2024

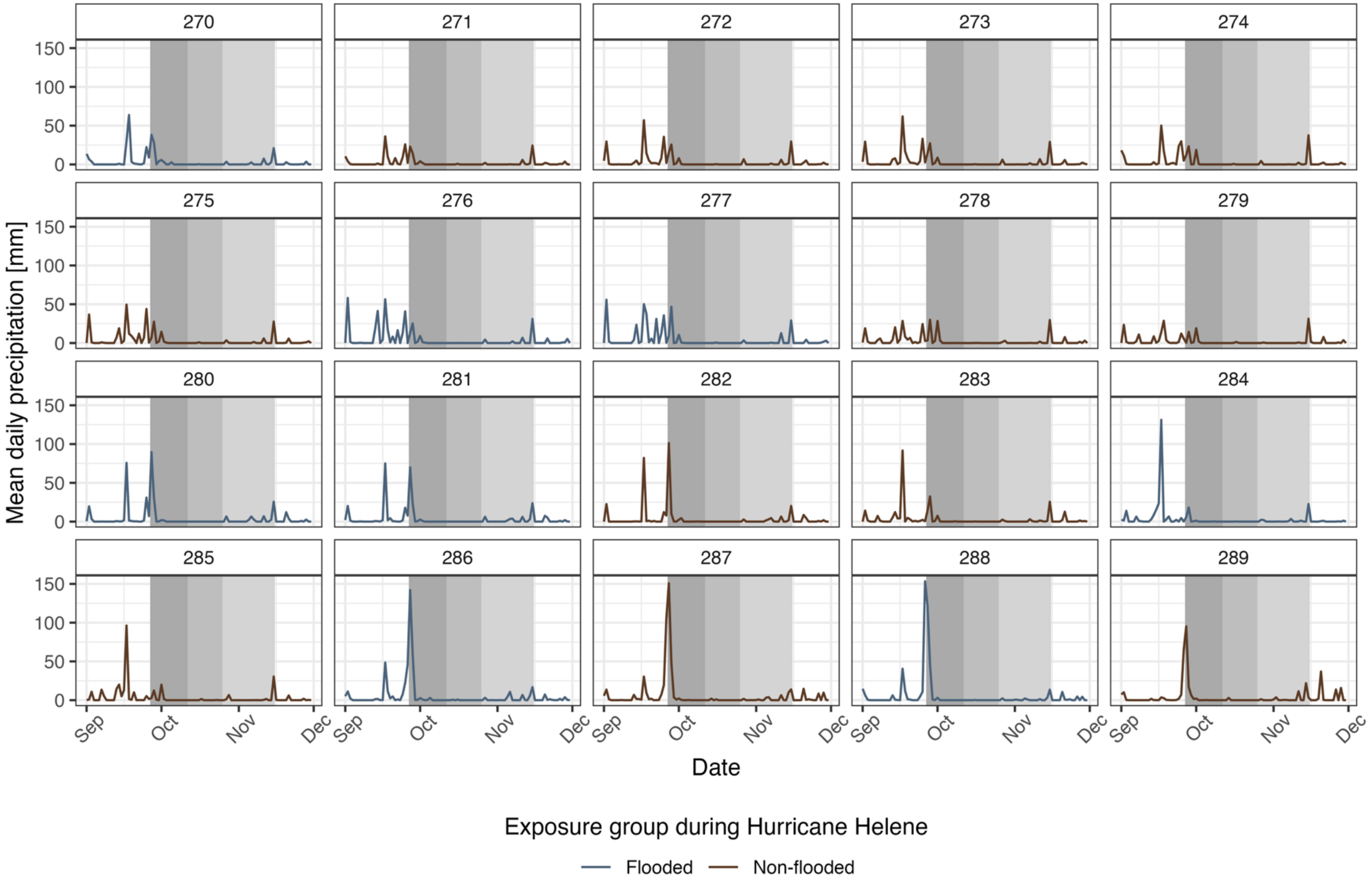


We obtained data on daily precipitation from the Parameter-elevation Regressions on Independent Slopes Model dataset.[5] Mean precipitation (shown in millimeters [mm]) was aggregated to the zip3-by-week level, but is shown above at the day-by-zip3 level to provide higher temporal resolution. Zip3 codes are shown at the top of each individual plot and the three-, five-, and eight-week periods following Hurricane Helene are shown with grey shading.

**Figure S 5:** Zip3 weekly trends in AGI visits (per 10k) from June to December 2024

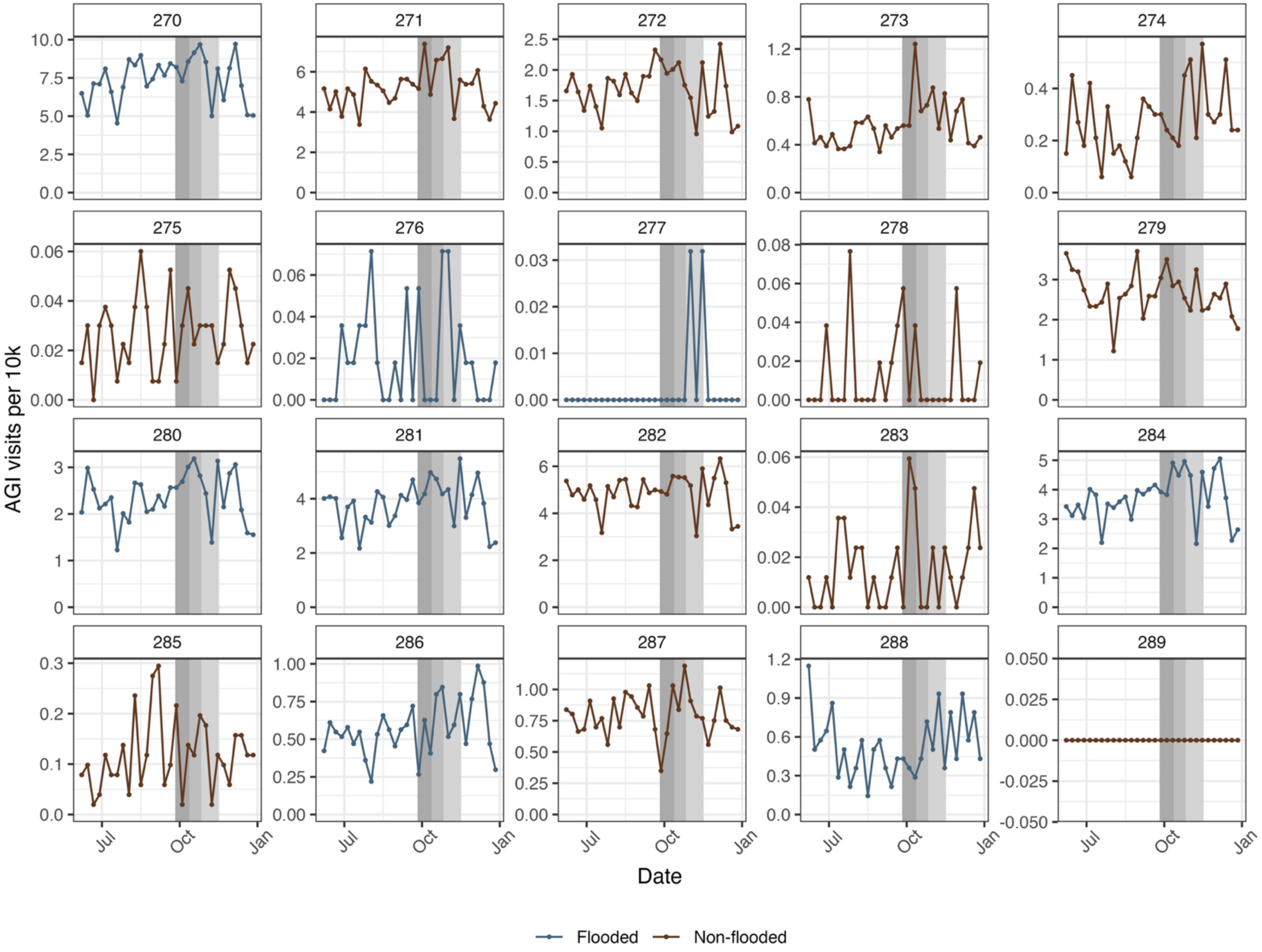


194 Zip3 codes are shown at the top of each individual plot and the three-, five-, and eight-week periods following Hurricane Helene are
195 shown with grey shading.

**Figure S 6:** Zip3-by-week level trends in AGI visits (per 10k) in the imputed datasets ($m$ = 20) from June to December 2024

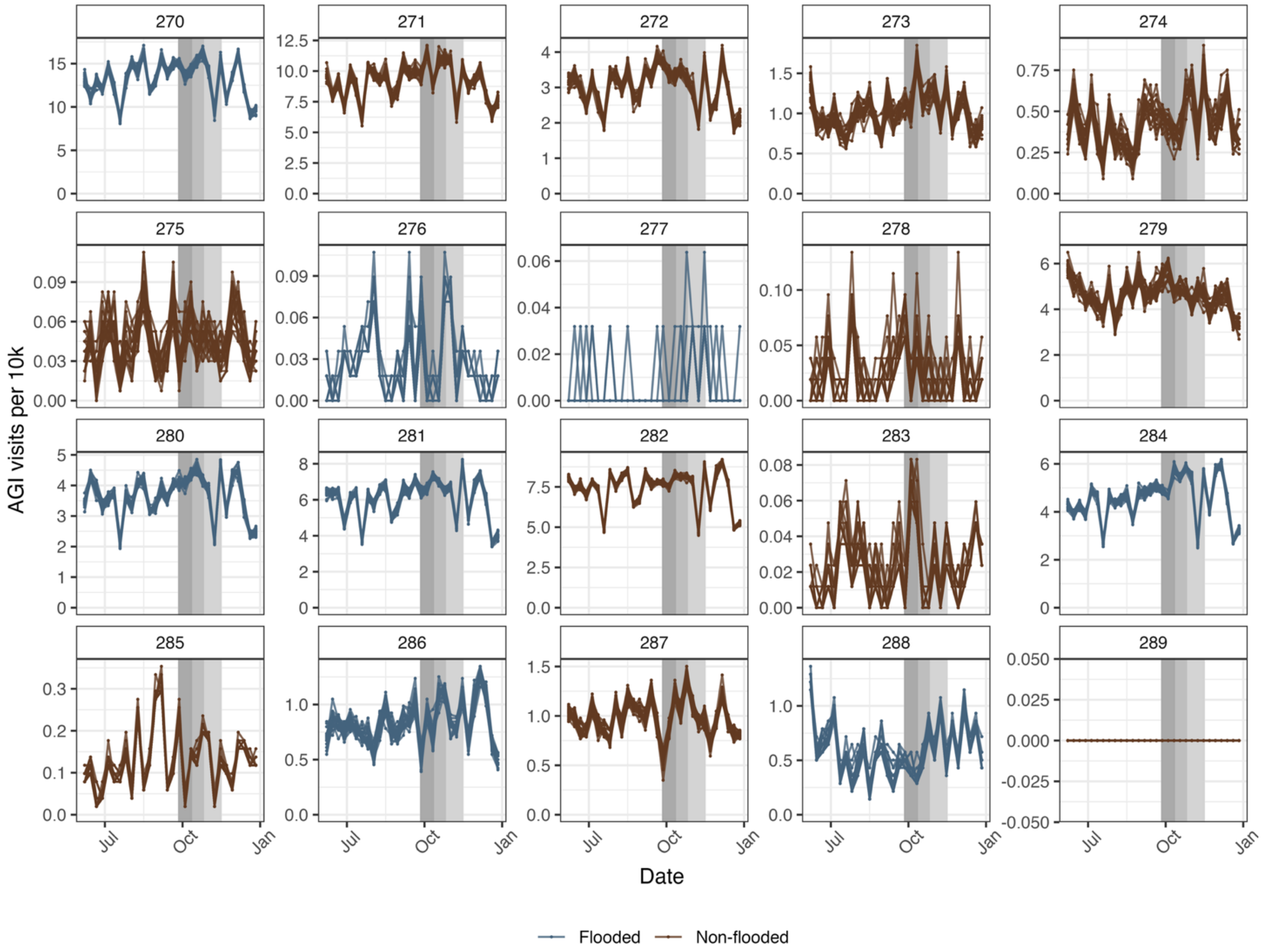


Zip3 codes are shown at the top of each individual plot and the three-, five-, and eight-week periods following Hurricane Helene are shown with grey shading.

### References


(1) US Census Bureau. *American Community Survey Data*. Census.gov. https://www.census.gov/programs-surveys/acs/data.html.

(2) US Census Bureau. *2020 Census Demographic and Housing Characteristics File (DHC)*. Census.gov. https://www.census.gov/data/tables/2023/dec/2020-census-dhc.html (accessed 2026-10-28).

(3) Murray, A.; Hall, A.; Riveros-Iregui, D. A Machine Learning Approach to Estimate Domestic Use of Public and Private Water Sources in the United States. *Water Research* **2025**, *276*, 123171. https://doi.org/10.1016/j.watres.2025.123171.

(4) *Global Flood Monitoring System (GFMS)*. http://flood.umd.edu/ (accessed 2026-07-28).

(5) *PRISM Group at Oregon State University*. https://prism.oregonstate.edu/data/ (accessed 2026-07-28).
